# Expanding Capabilities to Evaluate Readiness for Return to Duty after mTBI: The CAMP Study Protocol

**DOI:** 10.1101/2022.06.06.22276042

**Authors:** Amy Seal Cecchini, Karen McCulloch, Courtney Harrison, Oleg Favorov, Maria Davila, Wanqing Zhang, Julianna Prim, CDR Michael Krok

## Abstract

Physical Therapists affiliated with Intrepid Spirit Centers evaluate and treat Active Duty Service Members (ADSM) who have duty-limiting post-concussion symptoms to improve the ability to perform challenging tasks associated with military service. The Complex Assessment of Military Performance (CAMP) is a test battery that more closely approximates the occupational demands of ADSM without specific adherence to a particular branch of service or military occupational specialty. Subtasks were developed with military collaborators to include high level skills that all service members must be able to perform such as reacting quickly, maintaining visual stability while moving and changing positions, and scanning for, noting, and/or remembering operationally relevant information under conditions of physical exertion.

**Objective:** The purpose of this observational longitudinal study is to: 1- validate each component of the 3-task CAMP test battery; 2- examine responsivenss of the measures to change after participation in Physical Therapy; 3- develop reference values for clinical interpretation; and 4 - develop materials for clinical dissemination. This ongoing multi-site study is currently funded through the CDMRP and has been approved by the Naval Medical Center Portsmouth IRB.

**Methods:** ADSM undergoing post-concussion rehabilitation at the Intrepid Spirit Centers will be tested within one week of their initial Physical Therapy evaluation and after completing Physical Therapy. Control participants will include males, females, and ADSM from the Special Operations community. Participants will complete an intake form that includes questions about demographics, military service, deployment and concussion history, and profile and duty status. Other measures include those that explore concussion symptoms, sleep quality, post-traumatic stress, and perceptions of resilience.

The CAMP includes three separate 10-15 minute tasks. Movement is recorded by wearable inertial sensors and heart rate variability is recorded with a POLAR10 monitor. The “Run-Roll” task requires rapid position changes, combat rolls and quick running forwards and backwards while carrying a simulated weapon. Visual stability before and after the task is also performed. The “Dual-Task Agility” task includes rapid running with and without a weighted vest and a working memory task. The “Patrol Exertion” task requires repeated stepping onto an exercise step while watching a virtual patrol video. Additional tasks include monitoring direction of travel, observing for signs of enemy presence, and reacting to multiple auditory signals embedded in the video.

**Discussion:** Measures that evaluate relevant skills are vital to support safe return to duty for ADSM who may be exposed to imminent danger as part of training or mission demands. The CAMP is designed to be an ecologically valid and clinically feasible assessment that may be more sensitive to capturing subtle impairments that impact duty performance as test skills are integrated into dual and multi-tasks that reflect occupational demands. Assessment results may serve as a more robust indicator of readiness for full return to duty after concussion.

## INTRODUCTION

### Need for Military Return to Duty Assessments

An estimated 10-15%, with some reports of up to 30%, of Service members (SMs) who sustain mild traumatic brain injury (mTBI) experience activity limiting symptoms beyond the acute recovery period^1-4^, often requiring continued medical management or rehabilitation intervention. Active duty personnel must return to high levels of physical performance, using sensory, cognitive, and behavioral domains at levels that exceed targets of measures validated for civilian and athletic populations. Concussion specific measures to monitor the impact of intervention and track outcomes are needed to evaluate the effectiveness of clinical practice and the impact on force readiness^5,6^. Reliance on symptom report to gauge recovery is insufficient, due to factors including personality, stress, fatigue, aspects of military culture, warrior ethos, desire for gain, and recovery expectations that all impact self-assessment^4-10^.

Return to duty (RTD) requires physical, sensory, and cognitive readiness to resume all duties associated with the military occupational specialty (MOS). A major challenge of RTD decision making after concussion is the ability to predict whether a soldier who demonstrates clinical recovery is able to safely, efficiently and effectively perform duty specific and mission essential tasks^5,6,11,12^, including high level training activities in preparation for tactical duty in complex forward environments. Factors historically associated with successful RTD include a resolution of symptoms and improvement of distinct cognitive, motor and sensory impairments to perform functional, warrior specific tasks^4-6^. Single domain mTBI assessments may not fully reveal deficits that emerge under real world demands including stress or exertion, or during tasks that incorporate dual or multitask conditions^4-6, 12^. Many clinical tests are not sensitive beyond the first week post-injury and do not address the layered cognitive, sensory, and physical demands typical of military skills at challenging levels that avoid ceiling effects^5,6,12,13^. Ideal measures for the military population must assess relevant skills, and provide reference values in order to establish a clear differentiation between “impairment” and “readiness”. Advances in neuroimaging and biotechnology have revealed asynchronous symptom, clinical, and physiological recovery timelines and indicate that neurophysiological vulnerability and recovery extends beyond symptom recovery^14^. Premature RTD exposes the warrior to repeat injury while the brain is continuing to recover which may result in prolonged symptoms, more persistent dysfunction, and increased risk of additional injury all of which potentially impact the individual, the unit, and the mission.

Current guidance (Department of Defense Instruction 6490.11) mandates that all SMs who have sustained a concussion be evaluated prior to RTD and stipulates that those who have persistent symptoms beyond the acute recovery phase or those who have had 3 or more concussions within a 12 month period must undergo a “functional assessment” prior to return to duty.^11^ Efforts to develop measures that will meet this requirement challenge physical, sensory and cognitive skills (dual and multi-task scenarios), have proven effective in discriminating between those with concussion and their non-injured peers^13,15-16^. The Complex Assessment of Military Performance (CAMP) extends this prior work with 3 performance based tasks that target concussion vulnerabilities using sensitive physiologic measures. The purpose of this manuscript is to describe the methods proposed for cross-sectional validation of the test battery, as well as pre-post testing in a cohort of SMs with mTBI to examine responsiveness.

Investigation of the relationship of CAMP test variables to readiness for RTD, judged by clinician providers, will serve as the gold standard criterion, and SM perception of readiness for RTD will also be examined.

### Concussion Impairments Targeted in CAMP

Individuals with concussion may present with an array of cognitive and sensorimotor symptoms including headache, dizziness, imbalance, nausea and vomiting, sleep disturbances, sensitivity to noise and light, slowed thinking and reaction time, memory problems, irritability, depression, and visual changes^6,8,12^. These symptoms may contribute to a range of functional impairments.

Dual-task testing that requires a concurrent cognitive challenge during a motor task^17-19^ or multi-task scenarios that embed multiple cognitive tasks during the performance of physical tests^20^ has shown promise in identifying performance deficits after mTBI in sports concussion and military populations. The use of observational measures have value, as they are simple to administer clinically, but cognitive deficits are not always easily detected without sensitive measurement. Use of instrumented measures of reaction time may provide insight into changes in information processing that reflect subtle cognitive impairment post-injury.

Individuals who sustain concussion from blast exposures may experience additional vestibular symptoms including vertigo, oscillopsia, and motion intolerance, and, when injury occurs during combat-related activities emotional symptoms and stress reactions may co-occur beyond what is typical of civilians with mTBI^21^. Vestibular symptoms may relate to difficulty with balance and sensory integration necessary for military training and operational activities, and may affect the ability to respond to environmental sensory inputs (vision, body position). The ability to process information quickly and respond appropriately, in situations where sensory information may be difficult to interpret, is important for military operations. In training and combat, the ability to quickly change positions, move while carrying a weapon, and maintain exertion while closely attending to environmental conditions are critical^13,15-16^.

Physical exertion is an inherent part of military service and requires autonomic regulation of the cardiovascular system to respond to transitions in body position and dynamic physical demands^22-27^. Following mTBI some individuals experience exertional intolerance that is theorized to involve the autonomic nervous system and is identified through heart rate monitoring and the analysis of heart rate variability^28-34^.

The CAMP tests will allow the clinician to assess common physical impairments that may occur after mTBI with activities that mimic military training. Tests will incorporate cognitive elements including working memory and reaction time in dual- or multitask scenarios. Physical tasks challenge the vestibular system with rapid changes in head and body position. Exertion is required with brief fast paced tasks and with longer duration aerobic exercise. A lack of ability to perform cognitive and motor skills in combination may adversely affect performance of complex and often dangerous duty responsibilities, creating additional risk for individuals, units and mission success^6,13^. Availability of challenging, valid and objective measures may identify subtle but critical changes in function after injury that can be the focus for rehabilitation efforts^6^.

## METHODS

### Objective / Aims

This study is intended to improve on current practices to create a military-specific, valid and reliable functional assessment battery that assesses multiple domains of known vulnerability after concussion, in order to better inform providers and leaders on SM return to duty (RTD) readiness. The overall objective is to validate measures from a complex physical test battery that requires high level mobility and exertion paired with objective measurement via wearable sensors to track movement characteristics and response to exercise. Within the test battery are cognitive challenges that require working memory, visual attention, and instrumented reaction time responses. We hypothesize that SM with mTBI on average will demonstrate reduced capacity in movement tasks, greater cognitive task decrements in dual and multi-task conditions, slower reaction time during exertion, reduced HF HRV (during and after exercise). We further hypothesize that individuals who are recommended for return to duty after completion of physical therapy will, on average, demonstrate performance across tasks that approximates healthy control values at post-testing.

Aim 1 focuses on establishing typical parameters for active duty service members on the CAMP test battery by gathering healthy control performance data that will be used to interpret performance of individuals tested with mTBI. Aim 2 focuses on determining the elements of the CAMP battery that demonstrate the greatest differences from standard performance and those that serve as strongest predictors of successful return to duty using a regression model by gathering data from concussed personnel. An additional project aim is to refine clinician-facing feedback algorithms and displays, integrating data analysis and performance assessment results based on peer control performance variables.

### Design

This is an observational research study that uses known groups to evaluate performance on the CAMP test battery with two specific phases: 1- Cross sectional study of SMs with mTBI and healthy control SM peers and 2- Longitudinal pre-post therapy testing of SMs with mTBI who initiate and complete a physical therapy intervention. The first phase will examine group differences and also allow collection of data that can be used to provide reference values to judge typical performance. HC participants will be retested a month after the initial test to examine test retest reliability and clarify any learning or practice effects associated with the test battery. For the second phase, SMs engaged in physical therapy will be tested at the initation of PT and the week following discharge. The PT intervention will not be controlled, but provides an opportunity to capture change to assess responsiveness of the test battery.

The study will be conducted at 3 Intrepid Spirit Centers (Fort Bragg, Joint Base Lewis-McChord, Camp Lejeune) and will compare performance data between SMs with mTBI and their healthy peers.

### Sample Size

To achieve 80% power for group comparisons in longitudinal data analysis with consideration for attrition, power analysis for a logistic regression was conducted using the guidelines established in Lipsey & Wilson (2001) and G*Power 3.1.9.2 software to determine a sufficient sample size using an alpha of 0.05, a power of 0.80, a medium effect size (odd ratio = 1.72), and two-tailed test. The other covariates are expected to have a low association with the CAMP index score that will be developed to represent performance on the test battery (R=0.20, R2=0.04). The desired sample size is 128 subjects for logistic regression analysis. Prior work suggests that subjects in the mTBI group may become lost to follow up while others may not be able to complete all test elements resulting in incomplete data. Therefore, sample size is inflated by 15% from the largest size derived from above steps resulting in target enrollment of approximately 160 subjects in the concussed group. Target enrollment suggests a total of 335 subjects (160 concussed, 175 control) resulting in adequate power for all planned analyses. Attrition in the control group is expected to be insignificant with minimal missing data due to an inability to complete all test components. We plan to calculate the true power of the study for detection of any eventual smaller difference in study outcomes.

### Participants

Active duty service members across service branches between 18 and 45 years of age will be recruited. Exclusion criteria for all participants include hearing and vision deficits that impact function, major medical or psychiatric conditions or diagnoses, confirmed pregnancy, inability to safely participate in tasks that require a moderate amount of exertion, documented activity or profile restrictions that make participation in study activities unsafe, and lifetime history of moderate, severe, or penetrating brain injury. Concussed participants will be deemed eligible if their most recent documented concussion is between 2 weeks and 2 years prior to enrollment and they are currently receiving Physical Therapy at an Intrepid Spirit Center. Control participants will be deemed eligible if they have not had a concussion within the past two years and do not have ongoing symptoms requiring medical intervention for more remote concussion history. Control participants will be recruited to represent a range of test ages (18-22, 23-27, 28-32, 33-37, 38-45) with 20 in each group (n=100) recruited from general ranks of the military. Additional sampling will include special operations qualified personnel (n=50), as well as female ADSM (n=25) to allow for appropriate test interpretation for these groups.

Concussed participants from all sites will be tested within the first week of beginning physical therapy to examine differences in CAMP performance compared with the control group and will be re-tested after completion of physical therapy to evaluate change in CAMP performance. Control participants will be tested twice, at least 4 weeks apart, to determine the extent that prior exposure or task familiarity contributes to practice or learning effects. This information will be used to aid interpretation of re-test scores.

### Demographics and Self-Report Measures

Demographic variables include: age, sex, military occupational specialty (MOS), duty status, rank, branch of service, years of service, profile status, history of concussion, and deployment history. Self-report measures for all participants will include a measure of post-traumatic stress (Post traumatic stress Check List-Civilian, PCL-C), an index of common concussion symptoms (Neurobehavior Symptom Inventory, NSI), sleep quality (Pittsburth Sleep Quality Index, PSQI), and pain (Defense and Veterans Pain Rating Scale, DVPRS)^35-38^. Concussed participants also complete self-report measures to characterize their impairment complaints which could potentially change as a result of physical therapy intervention (Headache Intensity Test, 6 item, HIT-6, Dizziness Handicap Inventory, DHI) and a measure of resilience (Connor-Davidson Resilience Scale, CDRS)^39-41^.

### Test Battery

The protocol aims to increase objective measurement of impairments known to be problematic post-concussion that affect physical function, including dynamic visual acuity, tested before and after a task that requires rapid movement transitions and may provoke vestibular impairment; exertional tasks that challenge physical activity aerobically or with use of a weighted vest; reaction time challenges integrated into a complex task; motion sensor tracking via accelerometry and gyroscopes; and physiologic responses to exertion measured via heart rate variability analyses.

The test battery is administered in an order that increases the cognitive and physical load sequentially, to initiate performance testing even if symptom burden or exercise intolerance is a significant problem, beginning with 1- run/roll task, 2- dual-task agility without and eventually with a weighted vest to simulate a fighting load, and 3- patrol exertion multi-task scenario that integrates reaction time testing. The sequence of testing is illustrated in Figure 1. Heart rate is monitored throughout the test sequence track heart rate variability during recovery in 3 minute rest periods between tasks. An exploratory analysis of HRV during the exertional tasks will also be conducted. Inertial sensors worn on the head (for the first task) and lumbar area (for all tasks) track movement more sensitively than observational measures alone. Cognitive challenges are introduced in the dual-task agility and patrol exertion task. Symptoms are monitored at the beginning of the test battery, between each task during a 5 minute rest period, and at the end of the test battery using a Likert Scale for symptom intensity. Symptoms include “headache”, “visual instability”, “dizziness”, “nausea”, “fogginess”, or “other” which can be individualized by the participant. A rating of perceived exertion is recorded at these times as well.

**Figure 1.**
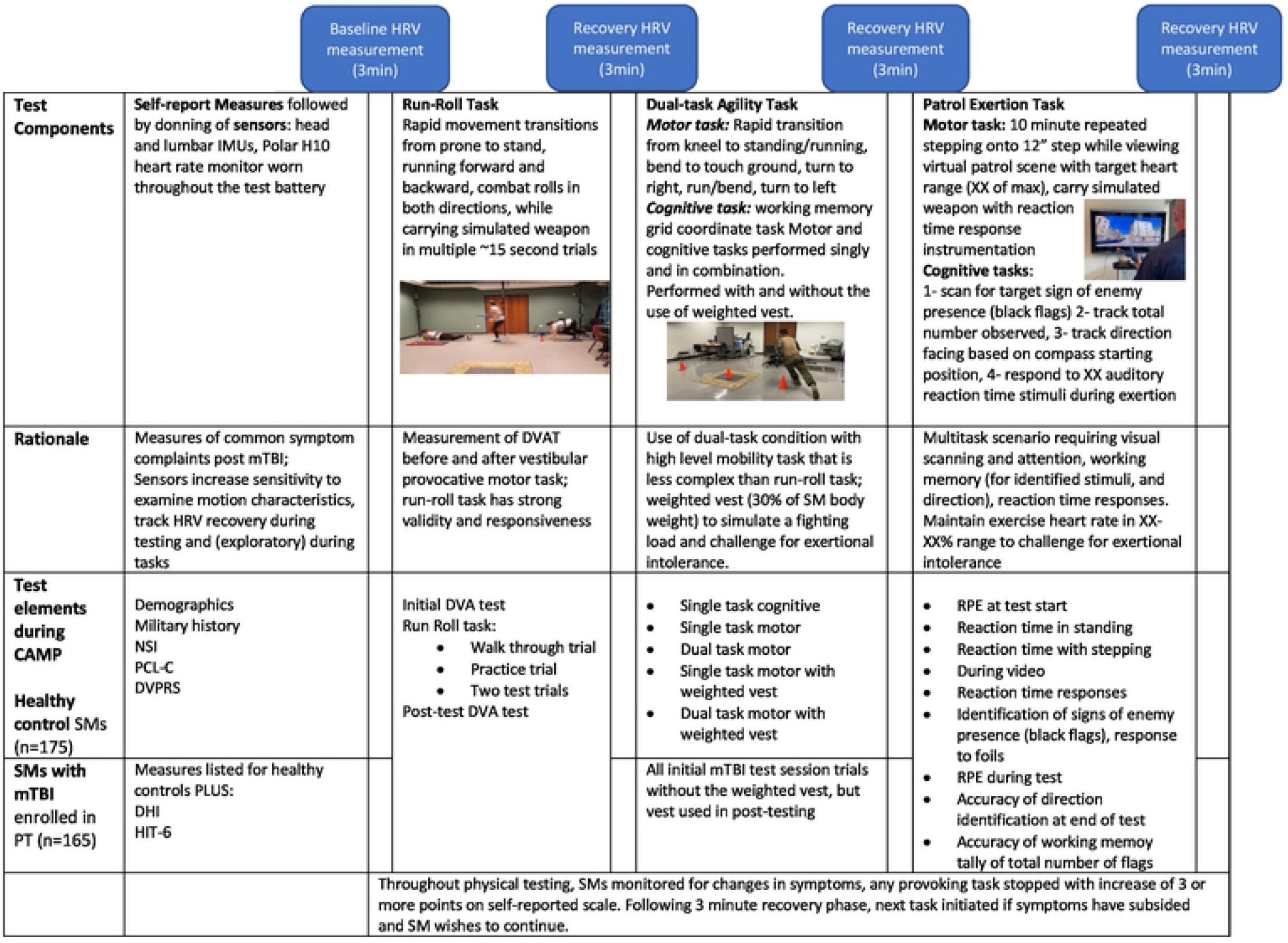
CAMP Test Battery Protocol.

A laptop receives multiple Bluetooth streams (reaction time, movement, HRV), collects sensor data, and in later phases of the project will automate signal processing and classification algorithms. The application has a simple user interface to guide the tester through the entire process. All of the testing components are turn-key systems that automatically connect to each other. Modular software architecture allows efficient upgrades and simplifies test administration. Extremely low power Bluetooth 4.0 communication allows for prolonged sensor life. High-bandwidth communication with the laptop is accomplished via a secure, mobile WiFi hotspot as public WiFi is often not available in military settings.

The entire CAMP test battery is administered using a program that synchronizes inertial sensor and heart rate data, aided by brief rest periods between tasks for heart rate recovery, allowing task components to be clearly identified for analysis.

### Task 1: Run/roll task

This task is based on the motor and visual stability components of the POWAR-TOTAL task^15^. The run/roll task demonstrated significant differences between ADSM post-concusion and their healthy control peers, and was responsive to change after completion of physical therapy^15,16^. The motor component is measured with tri-axial accelerometers and gyroscopes placed at the forehead, secured with a headband, and at the lumbar area, secured at the waist/belt line. Baseline seated dynamic visual acuity using a Snellen Chart and metronome to pace head turns at 120 Hz is conducted prior to the run/roll task^42^. The task begins in prone and requires carrying a simulated weapon. Once readiness is confirmed, trial start is triggered by a tone and vibration of the lumbar sensor that synchronizes trial initiation. The participant rapidly moves (1) prone to stand and a 10ft diagonal run, (2) stand-to-prone onto a floor mat and combat roll to the right, (3) prone-to-stand and run backwards to the start, (4) side shuffle to the left and diagonal 10 foot run to the mat, (5) stand to prone and combat roll to the left, (6) prone to stand and run backwards to the finish (Figure 2). The examiner manually indicates the end of each trial through the application on the computer. Each subject completes the task 4 times, with minimal rest between trials. The first trial allows practice and familiarization, subsequent trials are performed at the participant’s fastest pace. Immediately after all motor trials have been completed, the seated Dynamic Visual Acuity Test is repeated^43^.

**Figure 2.**
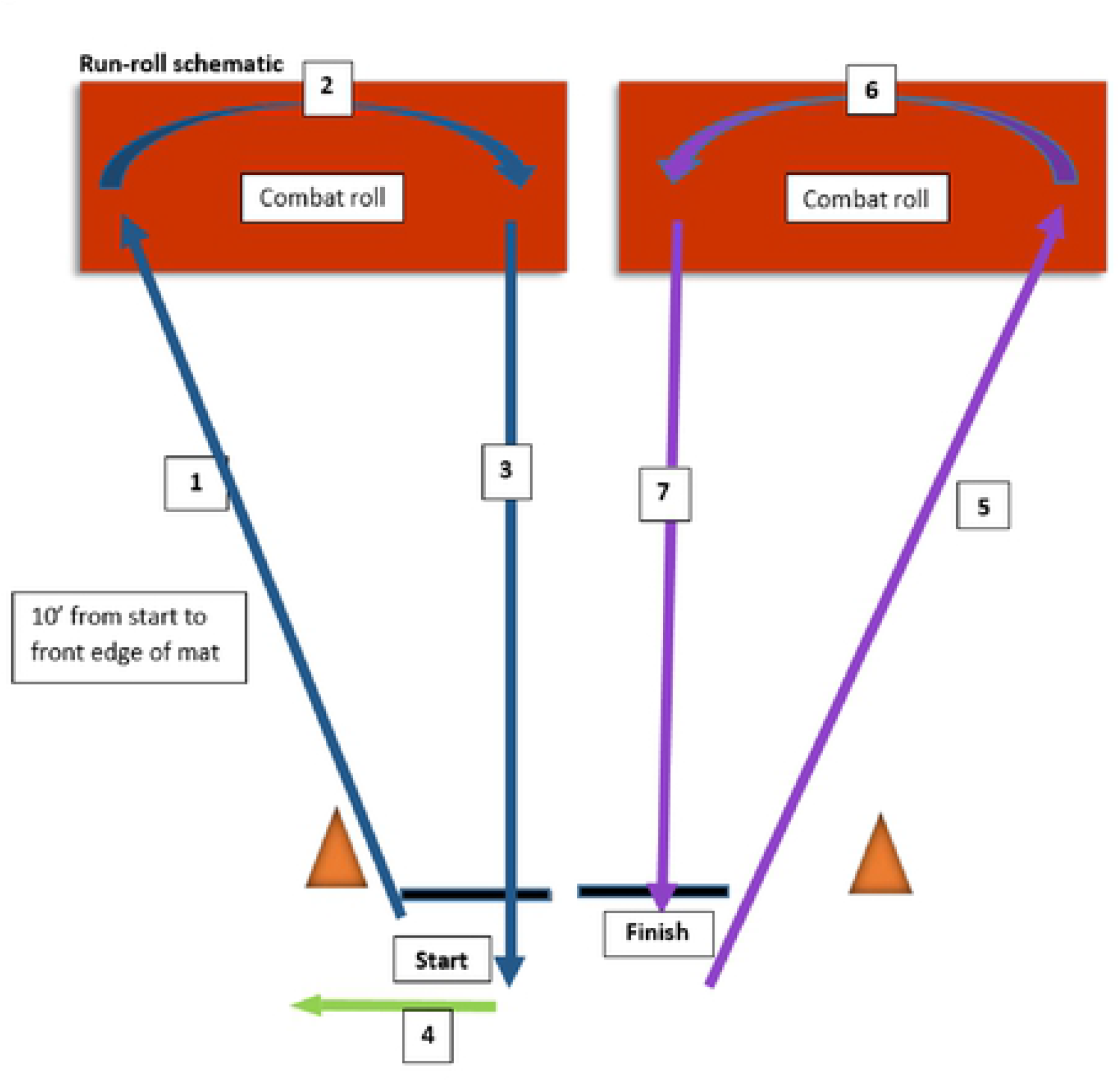
Run-Roll Task Sequence.

### TASK 2: Dual-task agility

Dual-task agility combines a motor and a cognitive task. A grid coordinate working memory task is used, as it is easy to administer and demonstrated between group discrimination ability in military mTBI studies^13,15^. ***Cognitive task:*** Participants begin with the cognitive task as a single task. An 8-character coordinate (2 alpha, 6 numeric) is read aloud to the participant, followed by a delay of 15 seconds. The participant is then asked to repeat the coordinate back in the exact order it was given, as best as they can recall. Scoring is based on stating the characters in the correct order. ***Motor task***: A hybrid of a shuttle run and the Illinois Agility Test, the motor task incorporates movement components that showed discriminative ability between healthy controls and concussed SMs in prior study^13^ but requires less space to allow its use in a typical clinic. Following familiarization with the task sequence (Figure 3), a single task motor trial is completed. The start position is in half kneeling. The participant runs forward 10 feet, touches a tape line on the floor with their left hand, turns inward and sprints back to the start, around a cone and then sprints forward 10 feet, touches a tape line with their right hand, turns inward, and sprints back to the finish line. The start of each trial is triggered by the beep and lumbar sensor vibration cue, with end of the trial marked by the examiner on the computer. Run time is recorded to the nearest hundredth of a second.

**Figure 3.**
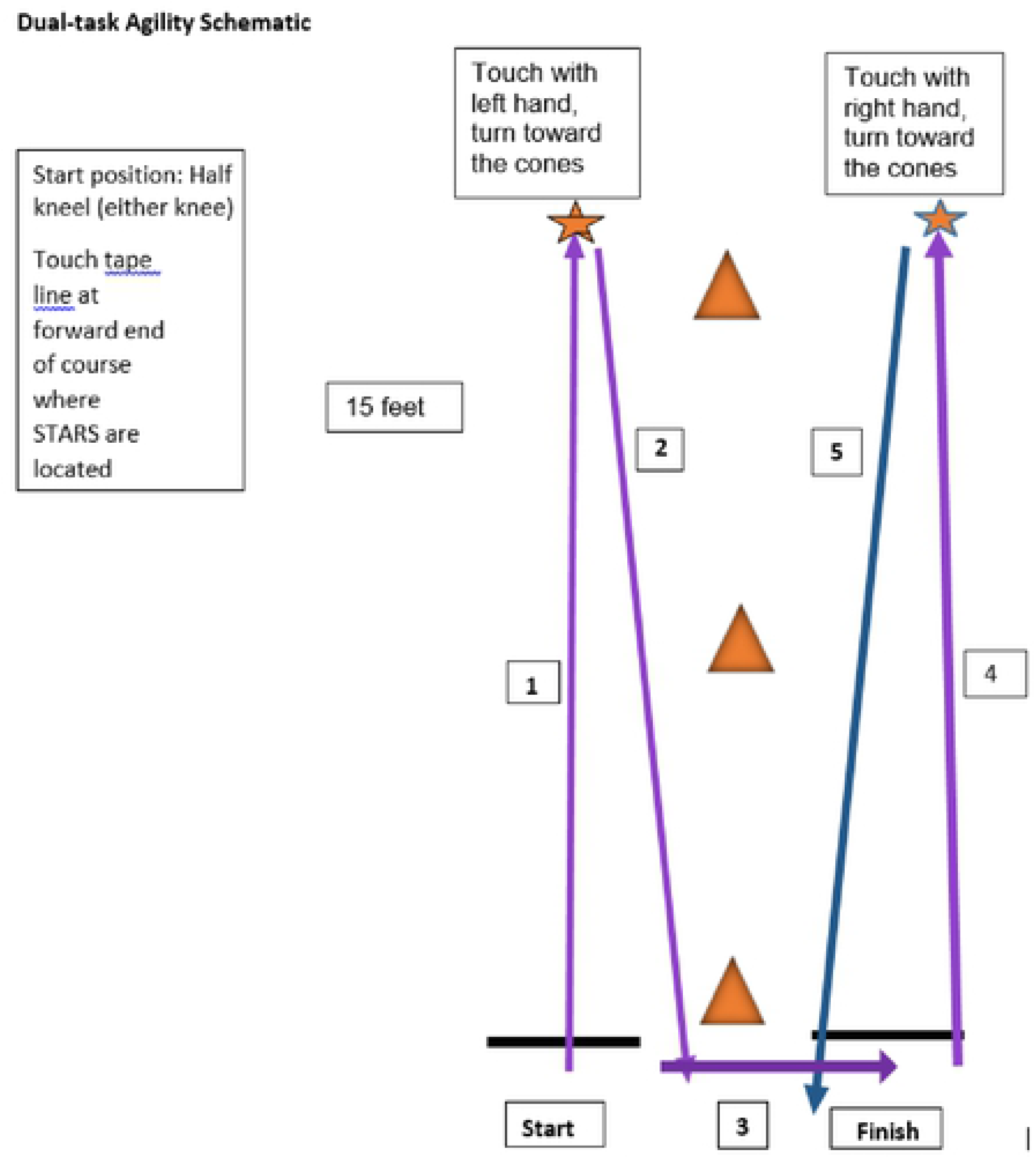
Dual-task Agility Sequence.

The task is then repeated in dual task conditions, where a similar but new grid coordinate is provided while the participant is in the start position. The course is run and the participant is asked to repeat the coordinate after crossing the finish line. Verbatim responses to the cognitive task are recorded manually and scored. A second single task motor trial is run followed by a second dual task trial. These repeat trials can be completed with the use of a weighted vest (30% of the participants’ body weight) in order to simulate the load that an ADSM might carry while wearing their body armor or other military gear. Control participants use the weighted vest in both initial and final test sessions. Concussed participants are tested without the weighted vest at the initial test session in order to avoid possible overexertion, but will use the weighted vest at the final test session, as after completion of their PT course they may be approaching clearance for return to full duty.

### Task 3: Patrol Exertion Task

#### Virtual Foot Patrol Scene

Two similar 10 minute virtual video files were created using a gaming platform to provide novel scenarios and allow for re-testing with reduced practice effects. A first-person camera angle was used to emulate visual scanning strategies and replicate the view experienced by an observer on a foot patrol in a village. The environment was augmented with a wide array of 3D models to incorporate unexpected but realistic audiovisual elements into the scene which would require the participants to attend and remain alert throughout the video. Elements incorporated in the scene were based on guidance from ADSM who have recovered from mTBI to improve task face validity. The scenes are displayed on a large monitor placed 4.5 feet in front of the participant at standing eye level to maximize visual immersion but accommodate the stepping task.

#### Motor Tasks

The participant carries a simulated M4 weapon (Bluegun^™^) equipped with a reaction time switch placed near the trigger. Participants are asked to continuously step up and down a 12” aerobic step while watching the virtual scene. The stepping pattern is selected by the service member, allowing for the leading limb to change during the test if they prefer. Exertion is targeted at a level consistent with 65-85% of age-based maximal heart rate^44^. The examiner interface displays the current heart rate with color coded indicators to ensure the appropriate exercise level (green – target range, yellow – heart rate is too low for target range, red – heart rate is too high for target range). Based on this feedback, the examiner provides cues to the participant: if yellow – step faster, if red – step slower. Ratings of perceived exertion are recorded at the beginning of the task and at the end of the task.

#### Multitask challenges

##### Reaction time

Instrumented reaction time data is collected throughout the patrol exertion task, in response to an auditory tone delivered via instrumentation on the weapon^45^. Initial responses to an auditory signal with two stimuli over a 20 second period are tested in standing to assure responses are sufficient (< 400ms), then in stepping without the video, and then for 12 stimuli interspersed at varying points in the video. The timestamp of the button press is recorded and sent to the laptop over Bluetooth. To ensure that the response time of a subject is accurately recorded, all devices are time-synchronized (nanosecond level) by sending a burst of time-stamped data packets from the instrumented weapon to the laptop at the beginning of the test. The laptop uses the timestamps and the inter-arrival time of consecutive packets in the burst to calculate the offset between the two clocks. Reaction time values are displayed via the user interface and recorded on the task scoresheet.

##### Additional cognitive elements

Additional task cognitive elements require visual attention, working memory and orientation to direction^46^. The participant is instructed to watch for and verbally identify specific targets of “enemy presence” (solid black flags) throughout the video that are presented at various locations. Participants verbally indicate each black flag they observe and keep a total count of the number of flags seen during the video. These responses are recorded manually by the examiner. Foil flags are also presented throughout the video. If these are identified by the participant, they are recorded. At the video start, a compass is embedded indicating the direction faced. The participant is advised they will need to report the direction they are facing at the end of the video. The video is then started. At the end of the video the participant is asked the direction they are facing and the number of signs of enemy presence they identified.

Outcome measurement from the CAMP test battery:

| **CAMP test item** | **Expected findings** | **Primary variables** | **Exploratory variables** |
| --- | --- | --- | --- |
| Run/roll task | More individuals with mTBI will be unable to complete the CAMP test components at initial examination (when beginning therapy). Individuals who are recommended for RTD will demonstrate movement “toward the healthy control” values | Duration of transitional movement (lowering from standing to prone, combat rolls) | Visual task post-testing<br>Head-torso coordination<br>Motor task time |
| Patrol Exertion task |  | RPE<br>Max exercise HR<br>Reaction time during task | Reaction time interindividual variability<br>Response to cognitive tasks |
| Dual-task agility task |  | Speed of turning during mobility task<br>Motor task time<br>Cognitive task accuracy | Dual-task interference |
| Heart rate variability collected during entire trial | Individuals with mTBI will have lower HRV, slower recovery of baseline HRV following tasks | RSA (HF-HRV) and Heart Period<br>Slope of HRV recovery during inter-task rest periods | LF HRV response during CAMP tasks (position changes in run/roll, exertion in patrol and dual-task agility) |

## DATA PROCESSING

### Inertial Sensor Analyses

Motor performance during the run/roll and agility tasks is recorded using inertial sensors attached to the body^16, 47^. Analyses will be performed on the time-series of tri-axial readings of the sensor’s accelerometer and gyroscope, sampled continuously at 100Hz during the task execution. Our previous POWAR-TOTAL study^16^ found that rapid transitional movements of lowering from running to prone position and of performing combat rolls were significantly more sensitive to mTBI than running. For example, according to our Receiver Operating Characteristic (ROC) curve analysis, SMs recovering from concussion are more accurately discriminated from their concussion-free peers based on the duration of the transitional movements (area under the curve AUC = 0.83) than on the duration of the entire task (AUC = 0.71).^16^This difference was hypothesized to be related to vestibular deficits in the concussed group. The greatest discriminatory difference was obtained on the last (fifth) trial. Therefore, in the CAMP study we will use the total time spent during the last trial while lowering from running to prone position and while rolling as our primary run/roll motor variable. In addition, we will also report the total duration of the last trial as our exploratory variable, since measuring the trial duration can be easily accomplished with a stopwatch and thus might be found more practical than inertial sensors in some clinical/operational settings.

To measure precisely the duration of lowering the body from the vertical to prone position and the duration of combat rolls, we will adopt the same machine-learning approach we used in the POWAR-TOTAL study, which involves training Radial Basis Function (RBF) networks on the accelerometer and gyroscope data recorded during run/roll activities^16^. During such training, individual RBF units become highly selectively tuned to different motor actions performed during the task, such as rising up from a prone position, running, lowering to prone, rolling, etc. The same approach will be used to measure the total duration and the speed of turning in the agility task.

Our previous POWAR-TOTAL study indicated that coordination of head and torso movements during the run/roll task is sensitive to mTBI and we will explore it in the CAMP study using Canonical Correlation Analysis. Coordination between the relative motions of the head and torso during run/roll task execution will be quantified by computing the full set of 6 canonical correlations between the coincident x, y, z values of the head and torso accelerometers and gyroscopes. The sum of squares of the canonical correlations will be used as an exploratory head-torso coordination metric of an individual’s motor performance.

### Heart Rate Variability Analyses

Heart Rate Variability (HRV) at rest has been tested extensively in military populations, but measuring HRV during exercise is less common. Baseline HRV and recovery of HRV will serve as our primary measure. We will explore HRV during the exertion elements of each task, but anticipate that the Patrol-Exertion task may be most useful given its longer duration and stability of body position. Heart rate and interbeat intervals (IBI) are recorded with the Polar H10 monitor wearable sensor. The Polar H10 has proven to capture heart rate with sufficient accuracy to derive HRV parameters^48^. Bluetooth transmission allows for direct transfer of the Polar H10 data to the laptop with time logs. Prior to analysis, each sequence of IBIs will be synchronized automatically, then manually inspected to ensure proper alignment of the IBI series with the timelog.

In this study we will be quantifying four parameters of HRV: RSA (high frequency HRV), heart period (HP), total HRV, and low frequency HRV (LF-HRV) to evaluate whether these additional indices provide insight into the changes in autonomic function during mTBI recovery. Beat-to-beat heart rate is continuously monitored (interbeat intervals IBI, time between consecutives R waves in milliseconds). Parameters of heart rate variability (HRV) are quantified^49,50^ to evaluate changes in neural regulation before and after the physical tasks. The unedited IBI output will be visually inspected and edited offline with CardioEdit software (University of Illinois at Chicago, 2007) in the first year of the project, automating this process for immediate feedback once we confirm data obtained during exercise is valid. Editing consists of integer arithmetic (i.e., dividing intervals between heart beats when detections of R-wave from the ECG are missed or adding intervals when spuriously invalid detections occurred). The remaining normal IBIs will be used in HRV analyses of RSA, LF-HRV and HP calculated with CardioBatch Plus software (University of North Carolina, Chapel Hill, 2016). The CardioBatch software implements the Porges-Bohrer method^50^. RSA calculated using this method is neither moderated by respiration, nor influenced by nonstationarity, and reliably generates stronger effect sizes than other commonly used metrics of HF-HRV^50^.

### Reaction Time Analyses

Mean reaction time will be examined between groups comparing values in standing, initial stepping and during the patrol task video^20^. The stimuli presented during the patrol task video will be divided into two groups (first and second group of 6 stimuli) to examine possible learning or fatigue effects. Intra-individual variability of reaction time responses will be examined, as higher variability in reaction time within a session has been demonstrated in those with mTBI^51^.

### Data management

Observational data are recorded on hard copy scoresheets. De-identified time stamped files for the inertial sensor and Polar monitor files are generated with each test session. Research staff at each site enter the data from the scoresheets into a RedCap database housed at University of North Carolina at Chapel Hill. Access to the database is controlled via PI registered enrollment with specific data management roles assigned as appropriate. The database offers interactive data entry with real-time field validation, audit logs to record modifications, integrity checks, security (in logins, permissions based on need, and encryption), reporting, forms inventory, and exports to common statistical packages for analysis. Data integrity checks are performed by a second team member to confirm accuracy of data entered from hard copy forms. De-identified motion sensor, Polar H10 files and Labview files that provide time code and heart rate test phases (baseline, task, recovery phases) are shared via MicroSoft Teams and exported to a university based server.

### Examiners

Research staff administering the protocol who are hired specifically for this project are educated at least at the bachelor’s degree level and are selected based on prior research experience and training, familiarity with military culture, technical problem-solving abilities, and professionalism. Onboarding includes security clearance to allow access to military research and health record systems and to access office and clinical space in their respective Intrepid Spirit Centers. Protocol administration training is conducted to ensure that testing is administered in a consistent manner. A detailed script that provides examiner and participant instructions for each task is used to train all examiners. Troubleshooting tips for technology malfunctions are provided via a shared project website. UNC based investigators provide training and offer extensive practice in administering the protocol through observation and hands on administration of computer interface management, task setup and instructions, and entire sequence testing prior to testing subjects for data collection.

### Recruitment

Recruitment of healthy control participants occurs through collaboration with TBI Center of Excellence researchers or through identified military collaborators who support research, in addition to approved flyer and social media advertisement. Recruitment of individuals who are recovering from mTBI occurs through collaboration with military site PIs who are physical therapists. Individuals who have undergone a Physical Therapy evaluation and are participating in rehabilitation for persistent symptoms associated with concussion will be identified by clinic providers. A member of the research team will contact those identified to provide information about the study. As each examiner manages all aspects of the study at each site, they are not blinded to participant status (healthy control, mTBI, first or second test) during testing. All participants who test during non-duty hours are offered a nominal gift card incentive ($25) for each test session they complete.

### Safety considerations

All testing is conducted in a controlled, air-conditioned clinical environment. Participants are closely monitored in order to observe for possible adverse reaction or symptom exacerbation while they are performing the tasks. Participants and test administrators will be wearing a mask during all study procedures in compliance with local, DoD, and CDC guidance regarding COVID-19 safety recommendations. Mask use during testing is recorded during each test session, to ensure initial test conditions can be replicated in post-testing. Tests done with and without masks will also be compared for possible differences in performance.

### Data and Statistical Analysis Plans

All statistical analyses will be performed using the SAS (Cary, NC) software package and will be performed consistent with clinically significant change comparing mean changes from baseline to post-intervention, and quantifying the magnitude of group differences between mTBI and healthy control groups. Effect sizes will be interpreted using Cohen’s guidelines (t-statistics: 0.2-0.4, 0.4-0.8, and > 0.8 representing small, medium and large effect size; f-statistics: 0.10, 0.25, and 0.40 represent small, medium, and large effect sizes, respectively). Preliminary assumption testing will be conducted to check normality, linearity, outliers, homogeneity of variance and multi-collinearity.

CAMP battery test performance will be analyzed for group differences using initial test session data. For each measure in the complete CAMP battery, a natural log transformation will be applied when a variable shows significant skew. Parametric data will be analyzed using the 2-tailed independent- sample t test to compare physiologic and functional differences between groups for each CAMP measure. Non-parametric data will be compared between the two groups using Chi-square. All other demographic and self-report measures will also be compared between groups which may reveal considerable variability. Further between group comparison between the concussed group final test measures and control group will allow for comparison of movement toward control values after physical therapy and will include the agility task with weighted vest comparison. Finally, multivariate models will be used to assess CAMP variables between the groups while controlling for variables that show significant differences at baseline, and which will be included as a covariant in the multivariate models.

Pre- and post-intervention assessments for the concussed group will also be performed to describe group demographics and clinical characteristics at baseline using stem-and leaf plots, frequency distributions and measures of central tendency and variability for each variable. Repeated measures ANOVA for testing whether means of CAMP test measures differ between tasks will be performed. For correlations between measures from the dual-task agility test (e.g., cognitive task), exertional test components (e.g., reaction time, HRV) and Run-Roll test (e.g., duration of transitional movements) Pearson correlation coefficients will be used for normal distributions and Spearman’s correlation will be used for non-normal distributions. A composite CAMP score will be formulated based on the above results by specific task measures. The concussed group will be divided into 3 subgroups based on the scores: high, low and non-responders. We will then include multiple comparison of pairwise means for testing whether means of CAMP components differ between the subgroups.

A regression model will be used to relate CAMP metrics at pre- and post-rehab time points to return to duty status. We will assess the relationship between overall CAMP measures and RTD after accounting for important covariates. We will use logistic regression where the outcome is a binary variable for RTD and the main predictor is the CAMP measure in mTBI group. RTD is not a simple construct, therefore RTD will be defined for this study as a full return to duty with all concussion related profile and duty restrictions being removed. The control variables include both demographic and clinical characteristics. Transformation will be conducted when appropriate to make the data normally distributed. The main predictor of composite CAMP index score is a continuous variable, and our regression model does not include multiple comparisons of pairwise means. Statistical analysis will include the following:

*First*, we will focus on assessing the independent CAMP measures (e.g., physiologic measures, functional measures, exertional test elements) for successful RTD outcome. Analysis by specific task measures will be conducted to examine any outcome differences between subjects with or without successful RTD outcome. *Second*, we expect that those characterized as high-responders to clinical intervention will show significantly different responses in composite CAMP measures than those who are low-responders or non-responders, providing validation for performance on CAMP battery as a predictor of RTD indicator. *Third*, there may be considerable baseline variability in clinical characteristics such as those measured by the Pain Rating Scale, Neurobehavioral Symptom Inventory (NSI) and possible medical comorbidities, and they can change over time, with or without an intervention. The possible medical comorbidities include headache, post-traumatic stress, and depression/anxiety disorders. A single numeric comorbidity sum score (combining these conditions) will be created. We will initially use the paired sample t-test to determine whether mean scores of these clinical variables differ before- and after-intervention. A univariate analysis will be conducted to identify these possible changes between pre- and post-intervention that might act as potential confounders on the association between composite CAMP measures and RTD. *Finally*, variables associated with successful RTD will be examined using both bivariate and multivariate analyses. The probability of RTD will be defined as the outcome variable and the composite COMP index will be used as the main predictor. Because the outcome variable RTD is binary (Pass/Fail), logistic regression models will be used with Wald tests to determine the significance of model covariates. Initial univariate analysis will be carried out by a χ2 test and a t-test to examine the relationship between outcome and the predictor variables, with statistically significant difference set at p < 0.05. We will also control for the demographic and additional independent variables that may be associated with the outcomes. These variables will include age, sex, readiness to deploy, MOS, total comorbidity scores, and the change in specific clinical characteristics (e.g., composite pain scale) at pre- and post-intervention based on the findings from the initial univariate analysis. Subsequent multivariable logistic regression with backward step entry will be applied; starting with a full model of all variables, a stepwise analysis will be used to remove non-significant variables one at a time to determine the independent predictors of outcome.

### Ethical Considerations

All appropriate legal and ethical considerations have been reviewed for the protection of human rights for voluntary participation by active duty service members. This study has been approved by Womack Army Medical Center, Madigan Army Medical Center, Navy Medical Center Camp Lejeune Human Protections Administrators and the Navy Medical Center Portsmouth (NCMP) Institutional Review Board (IRB). University of North Carolina IRB approval is also in effect, with reliance on the NMCP IRB.

### Status and timeline of the study

The study is planned for three years to include 2 years of active data collection, however the initiation of data collection was significantly delayed by the COVID pandemic. This study is currently open to enrollment at all 3 study sites.

## DISCUSSION

### Limitations of Study Design

The inclusion of healthy control participants who have a history of concussion is a limitation of this study. Many active duty roles are inherently risky, such that with increasing years of service, the likelihood that one may sustain mTBI increases. Individuals in the healthy control cohort are the peers of individuals who are injured. The ability to find a control group of “healthy” service members who range in age and years in service similar to that of the injured group and who have no history of injury is difficult. Our criteria therefore focused on those who are symptom free in spite of possible remote mTBI history and who are able to perform required duties without restriction. Analysis will include comparison of outcomes between those with and without concussion history to examine possible differences in our measures.

Examiners are not blinded to group or test session status which could result in bias, however many of our measures are collected via instrumentation or with computer based metrics that increase objectivity. The individuals with mTBI who are enrolled in our study participate in physical therapy, but that therapy course is not controlled, creating varied timelines between initial and post-testing. Given that the CAMP test battery is designed to aid in return to duty assessment, the benefit of testing individuals when they have met their PT goals and presumably are approaching duty readiness is a reasonable tradeoff. It is possible that differences in performance between those with mTBI and healthy controls in our test battery occur because of factors besides mTBI, including behavioral health conditions, pain, lack of conditioning, use of medications, or other factors that we do not measure. We do not envision the CAMP test battery as a diagnostic tool for TBI, rather as a mechanism to identify possible impairments that could affect RTD, whatever the cause, that will also be useful to track mTBI recovery. With additional study, it may prove useful for other groups as well.

### Plans for Dissemination

Early results that characterize the test-retest reliability of the CAMP battery for healthy controls will be shared, and determination of the need to continue re-testing for healthy controls will be made based on those results. We will share findings about our known group analyses and age band reference values as our sample sizes approach the required levels to complete these analyses. The pre-post therapy analyses will be disseminated later in the project as recruitment for those participants proceeds at a slower pace. We will share interim and final project findings at physical therapy, rehabilitation and military health conferences during the course of the project.

The grant proposal for this project included an aim to develop materials and resources to disseminate a validated CAMP test battery with TBICoE and other stakeholders. Ongoing input throughout the project will promote integration of findings with current guidance to improve concussion management in military populations and lay foundations for continuous product and resource development. We wil actively engage end users, service members and consultants regarding mechanisms to share test results in order to automate data analysis and provide feedback about SM performance soon after or during testing to the degree that is feasible.

## Conclusion

Concussion is common in active duty service members (ADSM) due to combat operations, but occures more often from garrison and community activities including airborne, weapons, artillery, and combative training; participation in sport; motor vehicle collision; or other accidental injury^1-3^. The CAMP test battery consists of ecologically valid activities relevant to military function as informed by military end-users. Clinicians will be able to use items from the test battery together or separately, focusing on common mTBI impairments in functional ways and at appropriate and individualized levels of challenge typical of the rehabilitation context. Flexibility to individualize the challenge will allow clinicians to identify impairments early in rehabilitation, while allowing for gradual additional difficulty to be added as rehabilitation progresses to approach the demands of active duty service, thereby informing return to duty decisions. The CAMP will be interpreted based on peer group reference data, providing immediate feedback for the therapist and patient.

The CAMP offers a feasible and clinician driven assessment that may provide a more comprehensive functional return-to-duty assessment protocol in order to identify tactically significant impairments for service members who would benefit from rehabilitation with the goal of reintegration and return to force capacity. Therapists working with concussed SM need a range of tasks with variable challenge capability to flexibly assess the physical, sensory, and cognitive challenges inherent in military duty. CAMP is intended to provide a standardized and validated test battery that is flexible to meet the needs of MTF clinicians who engage in return to duty decision making for SM diagnosed with concussion.

## Data Availability

Deidentified research data will be made publicly available through the FITBIR repository when the study is completed and published.

## Author Contributions

Amy S. Cecchini: conceptualization, writing – original draft; Karen L. McCulloch – conceptualization, supervision, project oversight, writing – original draft; Courtney Harrison – review and editing; Oleg Favorov – writing – review and editing, inertial sensor data collection and analysis plan; Maria Davila - writing – review and editing, heart rate variability data collection and analysis plan; Wanqing Zhang - writing – review and editing, data analysis plan; Julianna Prim - writing – review and editing, heart rate variability data collection and analysis plan; Michael Krok – local project administration, writing – review and editing.

## Acknowledgements

The CAMP team wishes to thank the active duty service members who havev contributed to the development of this project by participation in study of previous iterations of the test battery tasks and have participated in this project. We would not be able to conduct this study without our valuable project site principal investigators: CDR Michael Krok, Lisa O’Block, Christina Fitzpatrick, and LCDR Jonathan Gruber. Our CAMP project staff are instrumental in the project success: Annabell Oh, Carolyn Smith, Ashlee Bull, Hannah Neese, and Patricia Robinson. We also would iike to acknowledge UNC student physical therapists who havev assisted in the project: Michael Kress and Mikalia Guard.

